# Development and Evaluation of a Clinical Guideline for a Pediatric Telemedicine and Medication Delivery Service: A Prospective Cohort Study in Haiti

**DOI:** 10.1101/2023.02.15.23285858

**Authors:** Molly B. Klarman, Xiaofei Chi, Youseline Cajusma, Katelyn E. Flaherty, Anne Carine Capois, Michel Daryl Vladimir Dofiné, Lerby Exantus, Jason Friesen, Valery M. Beau de Rochars, Torben K. Becker, Chantale Baril, Matthew J. Gurka, Eric J. Nelson

## Abstract

**Objective:** Despite the emergence of telemedicine as an important model for healthcare delivery, there is a lack of evidence-based telemedicine guidelines, especially for resource-limited settings. We sought to develop and evaluate a guideline for a pediatric telemedicine and medication delivery service (TMDS).

**Methods:** A prospective cohort study was conducted at a TMDS in Haiti; children ≤10 years were enrolled. Among non-severe cases, paired virtual and in-person exams were conducted at the call center and household; severe cases were referred to the hospital. The primary outcome was the performance of the virtual exam compared to the in-person exam (reference standard).

**Findings:** A total of 391 cases were enrolled. Among 320 cases with paired exams, no general World Health Organization (WHO) danger signs were identified at the household; problem-specific danger signs were identified in 6 cases (2%). Cohen’s kappa for the designation of mild cases was 0.78 (95%CI 0.69-0.87). Among components of the virtual exam, the sensitivity and specificity of a reported fever were 91% (87%-96%) and 69% (62%-74%), respectively; the sensitivity and specificity of ‘fast breathing’ were 47% (21%-72%) and 89% (85%-94%), respectively. Kappa for dehydration assessments indicated moderate congruence (0.69; 95%CI 0.41-0.98). At 10 days, 95% (273) of the 287 cases reached were better/recovered.

**Conclusion:** This study, and resulting guideline, represents a formative step towards an evidence-based pediatric telemedicine guideline built on WHO clinical principles. In-person exams for select cases were important to address limitations with virtual exams and identify cases for escalation.

## INTRODUCTION

The provision of equitable healthcare has ascended to the forefront of the campaign to improve health outcomes globally. Target 3.8 of the United Nations Sustainable Development Goals (SDG) seeks to ‘Achieve universal health coverage’ and is foundational to attaining the other 12 health specific targets^1^. For example, access to timely treatment for childhood pneumonia is considered a key action to reduce preventable deaths of newborns and children under five (target 3.2). Healthcare coverage has increased, however gains have not been equitable, especially in resource-poor settings^2^. Following the current trajectory, coverage rates will likely fall short of targeted benchmarks^3^. Understanding who lacks access to healthcare, and why, is essential to meeting SDG 3.8. A framework used to study healthcare access barriers in low-income countries includes the following dimensions: geographical access, availability, affordability and acceptability^4^. Telemedicine is emerging as a model uniquely positioned to address challenges within each of these dimensions.

The COVID-19 pandemic was an accelerant for telemedicine adoption because it provided a mechanism to continue providing healthcare while minimizing risk of exposure^5^. Post-pandemic, telemedicine opportunities continue to emerge as a bypass to barriers limiting healthcare access^6,7^. Telemedicine can decompress workloads at clinics and emergency departments^8^, and increase access especially for rural^9^ and marginalized^10^ populations. There are multiple telemedicine models^11^. This study focuses on synchronous teleconsultations (telephone triage and advice)^8,12^ while drawing on community paramedicine models to extend care to households^13,14^.

Rapid telemedicine adoption is at risk of outpacing supporting evidence to assure safe and effective implementation. In 2010, the World Health Organization (WHO) identified telemedicine as a promising approach to equitably increase healthcare access as part of their Global Observatory for eHealth series^15^. In 2019, the WHO published the “Consolidated Telemedicine Implementation Guide” with a section on “Teleconsultation with Children and Adolescents”^16^. While comprehensive, the guidance focused on the environment for operating telemedicine services and did not include clinical guidelines. The lack of a telemedicine equivalent to the in-person WHO Integrated Management for Childhood Illness (IMCI) guidelines exposes a knowledge gap that must be addressed.

In response, we launched the Improving Nighttime Access to Care and Treatment (INACT) studies. The INACT1 study was a needs assessment in Haiti to characterize financial and logistical barriers to seeking care, and revealed telemedicine as a potential solution to bypass these barriers^17^. The findings were used to design a telemedicine and medication delivery service (TMDS) that targets the nighttime period when patients face the greatest barriers to access in-person care. The TMDS was piloted within the context of a prospective cohort study (INACT2); clinical safety and feasibility metrics have been described^18^. The objective of this study was to evaluate the pediatric telemedicine guideline that we derived from the WHO IMCI guidelines^19^ and implemented in this pilot. We hope this study inspires formative steps towards creating a telemedicine guideline that meets WHO standards. We also provide a framework to evaluate telemedicine guidelines using a virtual exam paired with an in-person exam as the reference standard.

## METHODS

### Ethics Statement

The study protocol was approved by the Institutional Review Board (IRB) at University of Florida (IRB201802920) and the Comité National de Bioéthique (National Bioethics Committee of Haiti; 1819-51). The study was registered at clinicaltrials.gov (NCT03943654).

### Study design

In this prospective cohort study at a TMDS in Haiti, we used paired virtual and in-person exams to evaluate the performance of a pediatric telemedicine clinical guideline.

### Study population and setting

The study took place in the commune of Gressier Haiti which has a population of approximately 38,100^20^. Gressier consists of both semi-urban and rural areas with agricultural and mountainous landscapes. Cellular coverage at the 3G level is sparse. There is no public electrical grid, household address system or streetlights. The TMDS operates with a delivery zone set to a 5 km radius (80 sq km) surrounding the call center. The under-five mortality rate is 63 per 1,000 live births; the global rate is 38 per 1,000 live births^21^. Leading causes of pediatric deaths are acute respiratory infection (ARI) and diarrheal disease^22^.

### Participant recruitment

Recruitment occurred through advertisement of the TMDS. Print and radio advertisements began two weeks prior to, and continued throughout, the study period.

### Participant inclusion criteria and consent process

Parent/guardians who contacted the TMDS during the hours of operation (6 pm - 5 am) about their child ≤10 years with a medical problem were eligible to participate. Written informed consent, and assent for participants ≥7 years, was performed upon household arrival. When no household visit occurred, the parent/guardian verbally agreed to a waiver of documentation of consent by phone.

### Enrollment estimates

The enrollment size for this prospective cohort study was based on a larger census-based prospective cohort pilot study described previously^18^.

### Participant incentives and fees

No incentives were offered. Families were informed that their willingness to participate in the study would not influence their ability to receive care. TMDS users were asked to pay a 500 Gourdes fee ($5US) to cover the medication and delivery cost. The fee was set on a sliding-scale down to zero to prevent the fee from acting as a barrier to accessing care. The fee served to assess willingness to pay, dissuade families from delaying seeking daytime care in favor of a free nighttime service, and to avoid conflict with fee-based daytime providers.

### Implementation

#### Staffing

The TMDS was staffed by licensed Haitian nurses/nurse-practitioners, motorcycle delivery drivers and on-call physicians. The physicians provided oversight, and situational adaptability for cases outside the scope of the clinical guideline.

#### Workflow

The INACT2 TMDS workflow (Figure 1) was described previously^18^. In brief, (i) A parent contacted the TMDS. (ii) A provider triaged the case as mild, moderate, or severe. (iii) Severe cases were referred to the hospital. (iii) For non-severe cases, a ‘virtual’ exam and medical history was performed to formulate an assessment and plan. (iv) For cases within the delivery zone, a driver and provider were dispatched to the household to conduct a paired in-person exam. (v) For cases outside the delivery zone, families received anticipatory guidance. Severe cases, and cases with a failed delivery, received a 24-hour follow-up call. All cases received a 10 day follow-up call.

**Figure 1.**
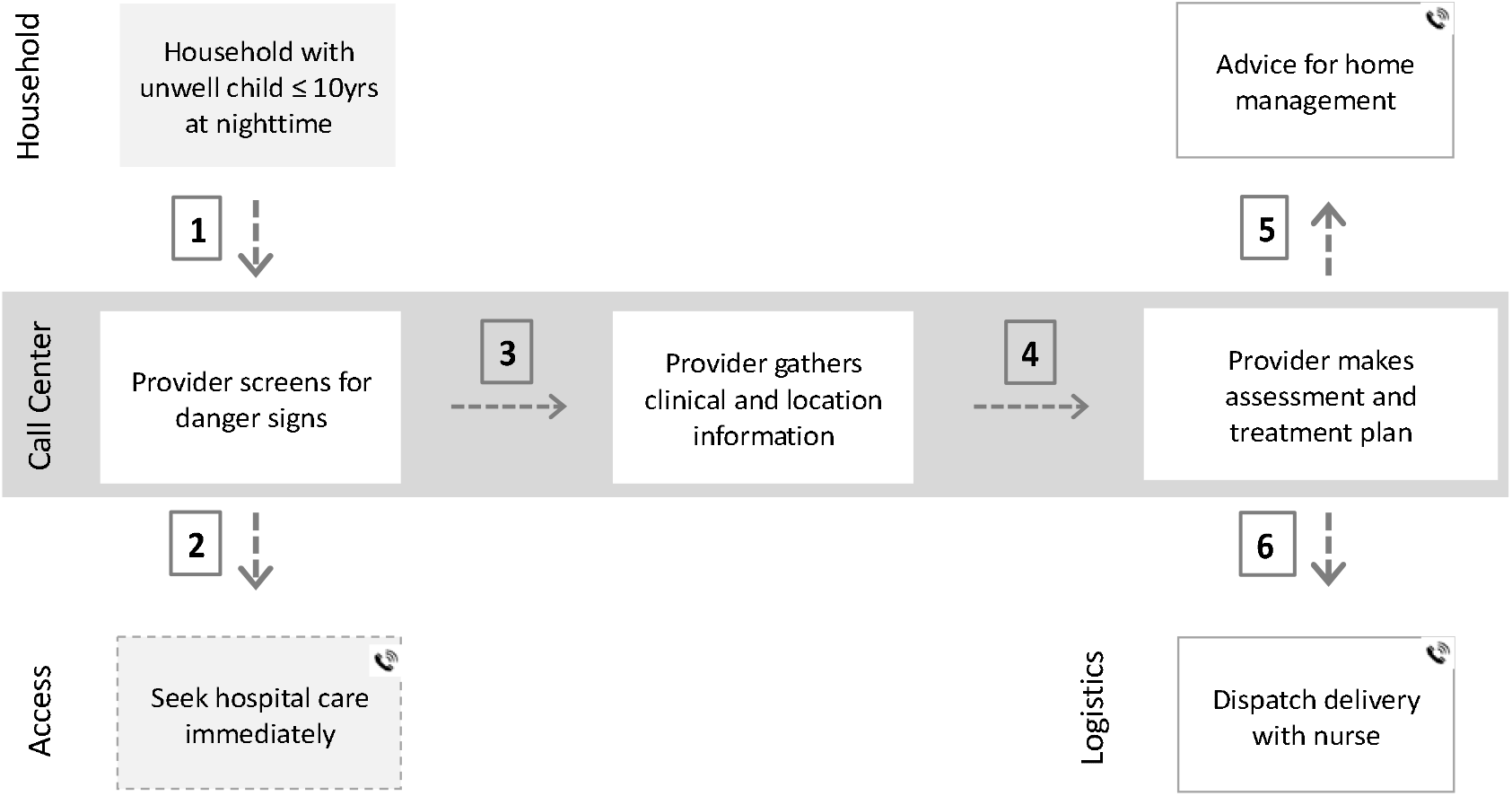
Telemedicine and medication delivery service workflow. (1) A parent/guardian contacts the call center. (2) A provider screens for danger signs; if present the child is referred to the hospital. (3) The provider gathers virtual exam findings and medical history. (4) An assessment and treatment plan are formulated. (5) If the child does not need medications/fluids or lives outside the delivery zone, the family receives anticipatory guidance alone. (6) If the child lives within the delivery zone and needs medications/fluids, a provider and driver are dispatched to conduct an in-person exam and transport items, respectively. Phone symbol = all families received a follow-up call at 10 days. A version of this figure has been published previously^1^ and is published here with permission from The American Journal of Tropical Medicine and Hygiene. Flaherty KE, Klarman MB, Cajusma Y, et al. A Nighttime Telemedicine and Medication Delivery Service to Avert Pediatric Emergencies in Haiti: An Exploratory Cost-Effectiveness Analysis. *The American Journal of Tropical Medicine and Hygiene*. 06 Apr. 2022 2022;106(4):1063-1071.

#### Clinical guideline and resources

The clinical guideline (Supplementary Text 1) was derived from the in-person WHO IMCI guidelines^19^, the WHO Integrated Management of Adolescent and Adult Illness (IMAI) guideline^23^ and the WHO Pocket Book for Hospital Care for Children^24^. Our conceptual model was to include only the most common medical problems based on our INACT1 needs assessment and WHO global burden of disease data^25^. These medical problems were: Fever, ‘Respiratory problem/Cough’, Dehydration/’Vomit’/Diarrhea, ‘Ear pain’, ‘Skin problem’, ‘Pain with urination’, and ‘Other’. The guideline for each problem consisted of an overview statement, diagnostic criteria based on the history and exam, criteria to triage cases as mild, moderate, or severe, and treatment location and follow-up recommendations. Recommendations were stratified by severity level. The guidelines provided the framework to navigate the case report form (CRF) which served as a paper clinical and logistical decision-support tool (Supplementary Text 2). A subset of questions on the CRF had fields for recording if the provider was ‘confident’ or ‘not confident’ in the response reported by the caller. This feature permitted situational adaptability in the case of a poor historian or technical challenges. A medication formulary with dosing recommendations was provided (Supplementary Text 3). The scope of use for these resources is restricted to this study. The intent is to iterate the materials within the INACT study series prior to generalized use, or consideration for adoption by the WHO.

#### Clinical procedures

Sample collection (stool, nasopharyngeal) and in-person measurements (vital signs and mid-upper arm circumference (MUAC)) have been described previously^26^.

#### Technology resources

Twilio Flex was used for call intake. Beacon software (Trek Medics International Inc.) was used to dispatch drivers and providers to households.

### Outcome measures

The primary outcome measure was the performance of the virtual exam compared to the in-person exam (reference standard). Data were analyzed by actionable domains within the clinical guidelines. The domains were triage level (mild, moderate, severe), WHO danger sign detection (problem agnostic and problem-specific), vital signs, and WHO assessment of dehydration (‘no’, ‘some’, ‘severe’). Secondary outcome measures included treatment plan adjustments after in-person examination, and clinical status at 10 days.

### Analytic strategy

The analysis was conducted according to the following framework: (i) A *post hoc* chart review, described previously^26^, was completed by two independent Haitian physicians to identify guideline deviations and if cases were ‘on’ or ‘off’ protocol. Variables that were ‘on protocol’ were analyzed. (ii) The first 5% of cases (washout period determined *post-hoc*) were excluded from the analyses, as were the second incidence of repeat participants within 30 days. (iii) Cases categorized as severe during the virtual exam and all other cases that did not receive a household visit were excluded from the analyses herein. (iv) Bacterial skin infection case severity was inadvertently miscategorized during implementation and was corrected *post hoc* (mild to moderate) prior to data analyses. (v) Responses marked as ‘not confident’ by the call center provider were not used for clinical decision making and excluded from the primary analyses. In a secondary analysis of the problem-specific respiratory and dehydration assessment questions, the ratios of the false positive to false negative responses were compared between ‘all’ and ‘confident only’ responses. An infant death with no causal relationship to the study occurred after the virtual exam and prior to the in-person exam was excluded from the paired analyses.

### Statistical analysis

Participant characteristics were described by proportions for categorical variables and medians for continuous variables. Binary assessments were described using Cohen’s kappa, sensitivity, and specificity, as well as positive predictive values (PPV) and negative predictive values (NPV). Assessments with kappa values of 0.01-0.20 were classified as no agreement, 0.21-0.39 minimal, 0.40-0.59 weak, 0.60-0.79 moderate, 0.80-0.90 strong, and >0.90 almost perfect^27^. We used the SAS ICC9 Macro to calculate the intraclass correlation coefficient (ICC) and 95% confidence intervals for congruence between continuous variables^28^. ICC values below 0.5 indicate poor agreement, while values 0.5-0.75, 0.76-0.9, and > 0.9 indicate moderate, good and excellent agreement, respectively^29^. Clustering effects from call center providers, repeat patient participants (>30 days between calls), or repeat adult parent/guardian callers were not considered. Analyses were completed using Statistical Analysis Software (SAS Institute) version 9.4.

## RESULTS

### Participant characteristics

Among 391 enrolled cases, 347 had paired virtual and in-person exams and 320 met criteria for analysis (Figure 2). The median age was 24 months, 48% (154) were <2 years, and 47% (150) were female (Table 1). The most common chief complaints were fever (44%; 142), ‘respiratory problem/cough’ (17%; 54), and ‘skin problem’ (15%; 49).

**Table 1.** Participant characteristics

| Characteristic | All cases included in analysis |
| --- | --- |
| All | 320<sup>a</sup> |
| Age; median mo (Q1-Q3)<sup>b</sup> | 24 (9-48) |
| <2 mo | 18 (6%) |
| 2 mo to <2 yrs | 136 (42%) |
| 2 yrs to <5 yrs | 105 (33%) |
| 5 yrs to 10 yrs | 61 (19%) |
| Sex; female | 150 (47%) |

<sup>a</sup> Enrolled cases that were excluded from analysis: no household visit (44), washout period (17), repeat patients within 30 days (9), severe adverse event prior to household visit (1).
<sup>b</sup> (Q1-Q3) = quartile 1 to quartile 3.

**Figure 2.**
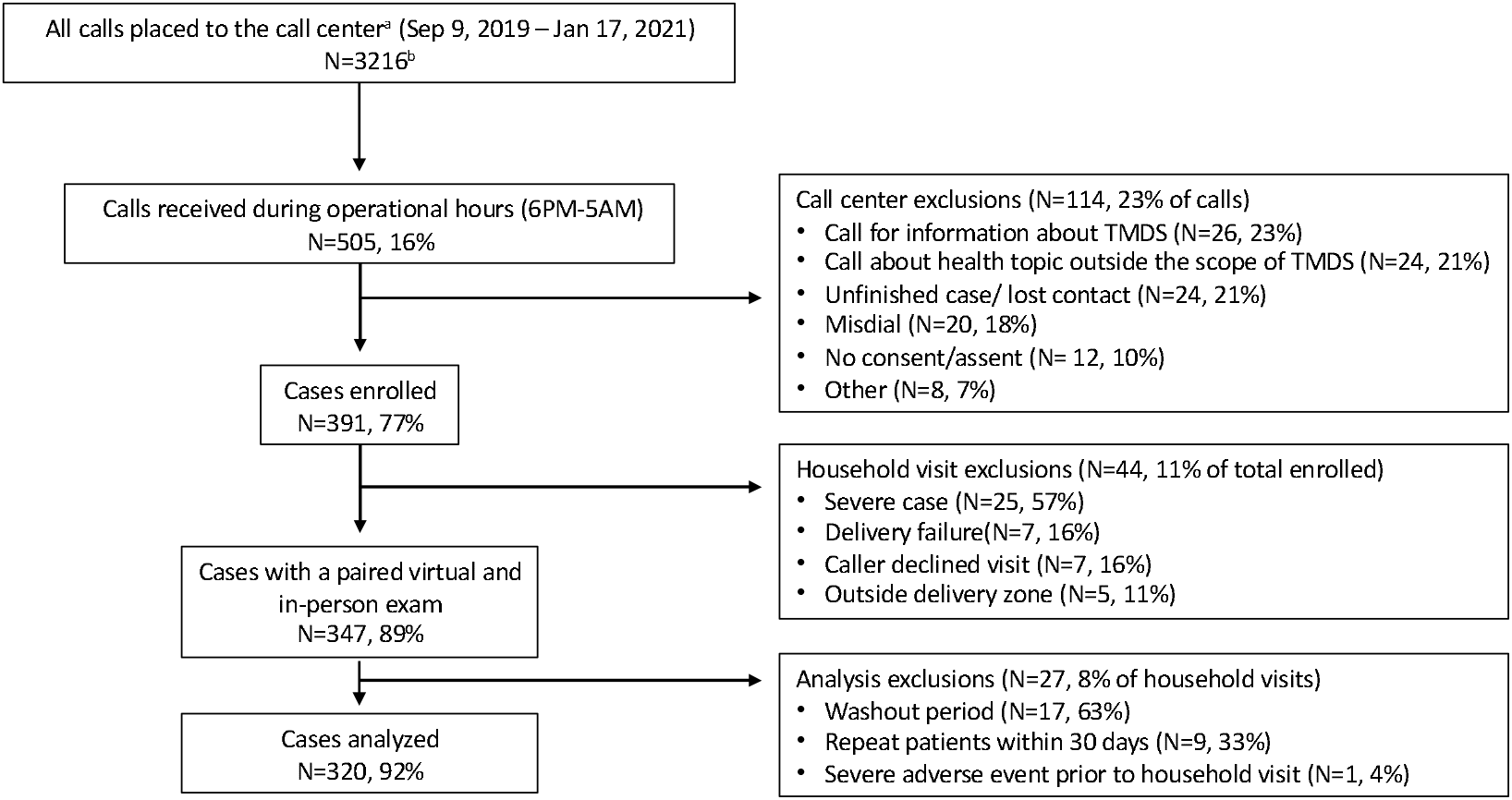
Diagram of case enrollment, reasons for exclusion and inclusion in data analysis. Of the 3216 calls placed to the call center during the study period, 505 callers were screened for inclusion. Among the 391 cases enrolled, 320 met criteria to be included in the analyses. ^a^ = filtered to remove calls from employee phone numbers. ^b^ = 1122 unique phone numbers.

### Primary outcomes

#### Triage level

The sensitivity of mild case severity assessed virtually was 95% (95%CI 93%-98%). The specificity was 83% (73%-93%) and the PPV and NPV were 96% (93%-98%) and 81% (71%-89%), respectively (Table 2). Cohen’s kappa indicated moderate agreement (0.78, 0.69-0.87). The sensitivity of moderate case severity assessed virtually was 86% (76%-95%). The specificity was 95% (92%-98%) and the PPV and NPV were 80% (69%-87%) and 97% (94%-98%), respectively. Cohen’s kappa also indicated moderate agreement (0.78, 0.69-0.87). Sub-analyses by disease type resulted in high levels of performance, with the exception of ARI with diarrhea.

**Table 2.** Performance of case severity determinations during the virtual exam

| Component | Severity | Total | CC+ HH+ (%)<sup>a</sup> | CC+ HH- (%)<sup>a</sup> | CC- HH+ (%)<sup>a</sup> | CC- HH- (%)<sup>a</sup> | Sens. (95% CI) | Spec. (95% CI) | PPV (95% CI) | NPV (95% CI) | kappa (95% CI) |
| --- | --- | --- | --- | --- | --- | --- | --- | --- | --- | --- | --- |
| All cases | Mild | 299 | 230 (77) | 10 (3) | 11 (4) | 48 (16) | 95 (93-98) | 83 (73-93) | 96 (93-98) | 81 (71-89) | 0.78 (0.69-0.87) |
|  | Moderate | 299 | 47 (16) | 12 (4) | 8 (3) | 232 (78) | 86 (76-95) | 95 (92-98) | 80 (69-87) | 97 (94-98) | 0.78 (0.69-0.87) |
| Cases by disease type |  |  |  |  |  |  |  |  |  |  |  |
| Fever w/o source<sup>b</sup> | Mild | 32 | 28 (88) | 1 (3) | 1 (3) | 2 (6.3) | 97 (90-100) | 67 (13-100) | 97 (85-99) | 67 (20-94) | 0.63 (0.16-1.00) |
|  | Moderate | 32 | 2 (6) | 1 (3) | 0 | 29 (91) | 100 | 97 (90-100) | 67 (23-93) | 100 | 0.78 (0.38-1.00) |
| ARI<sup>c</sup> | Mild | 75 | 63 (84) | 1 (1) | 2 (3) | 9 (12) | 97 (93-100) | 90 (71-100) | 99 (91-100) | 82 (53-95) | 0.83 (0.65-1.00) |
|  | Moderate | 75 | 8 (11) | 3 (4) | 0 | 64 (85) | 100 | 96 (91-100) | 73 (47-89) | 100 | 0.82 (0.62-1.00) |
| Diarrhea<sup>d</sup> | Mild | 42 | 33 (79) | 4 (10) | 0 | 5 (12) | 100 | 56 (23-88) | 89 (80-94) | 100 | 0.66 (0.37-0.96) |
|  | Moderate | 42 | 5 (12) | 0 | 9 (21) | 33 (79) | 56 (23-88) | 100 | 100 | 79 (71-84) | 0.66 (0.37-0.96) |
| ARI+Diarrhea | Mild | 10 | 7 (70) | 0 | 2 (20) | 1 (10) | 78 (50-100) | 100 | 100 | 33 (13-63) | 0.41 (-0.18-1.00) |
|  | Moderate | 10 | 1 (10) | 2 (20) | 0 | 7 (70) | 100 | 78 (50-100) | 33 (13-63) | 100 | 0.41 (-0.18-1.00) |

<sup>a</sup> CC= call center, HH= household, '+' = present, '-' = absent.
<sup>b</sup> Fever without source; acute respiratory infection (ARI), diarrhea, pain with urination, bacterial skin infection, scabies, vaginal discharge, and ear infections were considered medical problems with likely infectious etiology.
<sup>c</sup> ARI; cough with fever (excludes diarrhea).
<sup>d</sup> Diarrhea (excludes ARI).

#### Danger signs

Among the remaining cases that received in-person exams, no general WHO danger signs (lethargic/unconscious, seizure activity, unable to drink or breastfeed) were identified. Problem-specific danger signs were identified in 6 cases (2%) during the in-person exams.

#### Vital signs

Most fevers were reported subjectively (98%; 314). The virtual report of fever had a sensitivity of 91% (95%CI 87%-96%), specificity of 69% (62%-76%), PPV of 69% (62%-74%) and NPV of 91% (87%-96%) (Table 3). The categorization of fast vs. not fast breathing, per WHO cut-offs, had a sensitivity of 47% (21%-72%), specificity of 89% (85%-94%), PPV of 29% (11%-47%), and NPV of 95% (91%-98%). The continuous variable of respiratory rate had poor agreement (ICC 0.42; 95%CI 0.31-0.55) (Table 4) as did heart rate (ICC 0.27; 0.11-0.51).

**Table 3.**
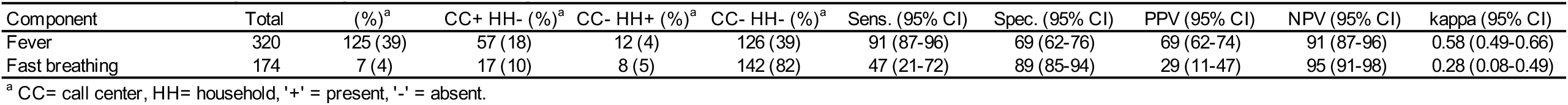
Performance of categorical vital sign determinations during the virtual exam

| Component | Total | (%)<sup>a</sup> | CC+ HH- (%)<sup>a</sup> | CC- HH+ (%)<sup>a</sup> | CC- HH- (%)<sup>a</sup> | Sens. (95% CI) | Spec. (95% CI) | PPV (95% CI) | NPV (95% CI) | kappa (95% CI) |
| --- | --- | --- | --- | --- | --- | --- | --- | --- | --- | --- |
| Fever | 320 | 125 (39) | 57 (18) | 12 (4) | 126 (39) | 91 (87-96) | 69 (62-76) | 69 (62-74) | 91 (87-96) | 0.58 (0.49-0.66) |
| Fast breathing | 174 | 7 (4) | 17 (10) | 8 (5) | 142 (82) | 47 (21-72) | 89 (85-94) | 29 (11-47) | 95 (91-98) | 0.28 (0.08-0.49) |

<sup>a</sup> CC= call center, HH= household, '+' = present, '-' = absent.

**Table 4.** Performance of continuous vital sign measurements during the virtual exam

| Component | ICC<sup>a</sup> (95% CI) |
| --- | --- |
| Respiration rate | 0.42 (0.31-0.55) |
| Heart rate | 0.27 (0.11-0.51) |

<sup>a</sup> ICC= interclass correlation coefficient.

#### Dehydration screen and assessment

The virtual dehydration assessment for ‘no’ dehydration had a sensitivity of 97% (95%CI 93%-100%), specificity of 83% (53%-100%), PPV of 99% (97%-100%) and NPV of 63% (29%-96%); Cohen’s kappa was 0.69 (95%CI 0.41-0.98). Of the 97 cases evaluated for dehydration, one instance of moderate dehydration was not detected by virtual exam. Individual components of the WHO dehydration assessment had variable performance and samples sizes for absence of urine or tears were too low to make statistical inference (Table S1).

### Secondary outcomes

Changes to the medication treatment plan after in-person exams were uncommon; Cohen’s kappa values were generally above 0.75 (Table 5). The in-person exams resulted in amoxicillin removal for 9% (26) of cases and addition of amoxicillin for 3% (8) of cases. At 10 days, 95% (273) of cases included in these analyses were better/recovered.

**Table 5.** Congruence of medications prescribed between the virtual and in-person exams

| Medication type<sup>b</sup> | Total | CC+ HH+ (%)<sup>a</sup> | CC+ HH- (%)<sup>a</sup> | CC- HH+ (%)<sup>a</sup> | CC- HH- (%)<sup>a</sup> | kappa (95% CI) |
| --- | --- | --- | --- | --- | --- | --- |
| Paracetamol | 305 | 168 (55) | 13 (4) | 9 (3) | 115 (38) | 0.85 (0.79-0.91) |
| Amoxicillin | 305 | 86 (28) | 26 (9) | 8 (3) | 185 (61) | 0.75 (0.68-0.83) |
| Cephalexin | 305 | 35 (12) | 5 (2) | 7 (2) | 258 (85) | 0.83 (0.74-0.92) |
| Benzyl benzoate | 305 | 30 (10) | 2 (1) | 2 (1) | 271 (89) | 0.93 (0.86-1.00) |
| Zinc | 305 | 18 (6) | 1 (<1) | 1 (<1) | 285 (93) | 0.94 (0.87-1.00) |
| Neomycin | 305 | 11 (4) | 1 (<1) | 3 (1) | 290 (95) | 0.84 (0.69-0.99) |

<sup>a</sup> Medications with ≥10 household prescriptions are included.
<sup>b</sup> CC= call center, HH= household, '+' = present, '-' = absent.

### Exploratory analyses

For the aggregate of four respiratory components, the responses that were marked as ‘confident’ had a ratio of false positive (19) to false negative (11) responses of 1.7; without this designation, the ratio was 19.3 (135/7) (Table 6). For the aggregate of the five dehydration assessment components, the responses that were marked as ‘confident’, had a ratio of false positive (14) to false negative (14) responses of 1.0; without this designation, the ratio was 2.0 (48/24) (Table 7).

**Table 6.** Evaluation of a ‘confidence’ score when assessing a breathing problem during the virtual exam

| Component | Independent of 'confidence' score<sup>a</sup> |  |  | Dependent on 'confidence' score<sup>b</sup> |  |  |
| --- | --- | --- | --- | --- | --- | --- |
|  | CC+ HH-<sup>c</sup> | CC- HH+<sup>c</sup> | Ratio False+<sup>d</sup>/False-<sup>e</sup> | CC+ HH-<sup>c</sup> | CC- HH+<sup>c</sup> | Ratio False+<sup>d</sup>/False-<sup>e</sup> |
| Head bobbing | 29 | 1 | 29.0 | 5 | 1 | 5.0 |
| Nasal flaring | 56 | 4 | 14.0 | 12 | 6 | 2.0 |
| Retractions | 35 | 2 | 17.5 | 2 | 3 | 0.7 |
| Stridor | 15 | 0 | - | 0 | 1 | 0.0 |
| Total | 135 | 7 | 19.3 | 19 | 11 | 1.7 |

<sup>a</sup> Data shown are from all respondents during the virtual exam, regardless of the provider's confidence in the response.
<sup>b</sup> Data shown are only for cases in which the provider was 'confident' in the response conveyed during the virtual exam.
<sup>c</sup> CC= call center, HH= household, '+' = present, '-' = absent.
<sup>d</sup> False + = The virtual assessment indicates a finding is present but during the in-person assessment it is absent.
<sup>e</sup> False - = The virtual assessment indicates a finding is absent but during the in-person assessment it is present.

**Table 7.** Evaluation of a ‘confidence’ score when assessing dehydration during the virtual exam

| Component | Analyses independent of 'confidence' score<sup>a</sup> |  |  | Analyses dependent on a 'confidence' score<sup>b</sup> |  |  |
| --- | --- | --- | --- | --- | --- | --- |
|  | CC+ HH-<sup>c</sup> (%) | CC- HH+<sup>c</sup> | Ratio False+<sup>d</sup>/False-<sup>e</sup> | CC+ HH-<sup>c</sup> | CC- HH+<sup>c</sup> | Ratio False+<sup>d</sup>/False-<sup>e</sup> |
| Disposition ≥5 yrs |  |  |  |  |  |  |
| Alert ≥5 yrs | 0 | 0 | - | 0 | 0 | - |
| Disposition <5 yrs |  |  |  |  |  |  |
| Alert <5 yrs | 1 | 6 | 0.2 | 2 | 3 | 0.7 |
| Irritable | 4 | 2 | 2.0 | 3 | 2 | 1.5 |
| Lethargic | 3 | 0 | - | 0 | 0 | - |
| Sunken eyes | 24 | 0 | - | 2 | 2 | 1.0 |
| Thirst |  |  |  |  |  |  |
| Normal | 1 | 8 | 0.1 | 1 | 3 | 0.3 |
| Thirsty | 7 | 1 | 7.0 | 3 | 1 | 3.0 |
| Unable to drink | 1 | 0 | - | 0 | 0 | - |
| Skin pinch |  |  |  |  |  |  |
| Normal | 1 | 6 | 0.2 | 1 | 2 | 0.5 |
| Slow | 5 | 1 | 5.0 | 2 | 1 | 2.0 |
| Very slow | 1 | 0 | - | 0 | 0 | - |
| Total | 48 | 24 | 2.0 | 14 | 14 | 1.0 |

<sup>a</sup> Data shown are from all respondents during the virtual exam, regardless of the provider's confidence in the response.
<sup>b</sup> Data shown are only for cases in which the provider was 'confident' in the response conveyed during the virtual exam.
<sup>c</sup> CC= call center, HH= household, '+' = present, '-' = absent.
<sup>d</sup> False + = The virtual assessment indicates a finding is present but during the in-person assessment it is absent.
<sup>e</sup> False - = The virtual assessment indicates a finding is absent but during the in-person assessment it is present.

## DISCUSSION

In this prospective cohort study, the performance of a pediatric telemedicine guideline was evaluated by comparing virtual and in-person exams. Cases without WHO general danger signs identified during the virtual exam, also had no danger signs identified during the in-person exam. However, two percent of cases had a problem-specific danger sign identified in-person. During the virtual exam, vital sign questions had mixed performance and dehydration assessments were reliable for identifying ‘no’ dehydration. After in-person exams, changes to medication treatment plans were uncommon and often resulted in improved antibiotic stewardship. A need for situational adaptability emerged as a critical aspect of the guideline development. These findings represent formative steps towards an evidence-based telemedicine guideline for resource-limited settings.

The WHO framework for in-person triage classifies cases as severe, moderate, or mild. The approach uses general and problem-specific danger signs. Our research challenge was to test the hypothesis that aspects of the WHO framework could be adapted for use in a virtual telemedicine environment. We expanded the triage process to include logistical constraints that would require escalated care (e.g., need for nebulized salbutamol). The findings suggests that the guideline was accurate at identifying mild cases. There was a low, yet meaningful, discovery rate of cases with problem-specific danger signs. Therefore, in-person examinations are required for select cases, especially for moderate cases that are at risk of converting to a severe status. These results are consistent with other telemedicine studies on triage^30^.

Our study was designed to test the limits of telemedicine at households with limited connectivity, electricity, and medical knowledge. Callers were asked to report fever and count breaths and heart beats. The results suggest that future iterations of the guideline should continue to inquire about fever and respiratory rate, but no longer include heart rate. While the assessment of dehydration identified cases with no dehydration, the virtual exam failed to accurately identify patients with sunken eyes (one of the four WHO dehydration assessment components). This result demonstrates that not all components of the WHO in-person guidelines can be transferred to a virtual telemedicine guideline. A new scoring system may be needed that leverages those components of the virtual exam that are reliable and discards those deemed unreliable.

Exploration of individual responses was required. Asking questions ‘around’ the primary questions exposed important insights (e.g., paracetamol ingestion) that might confound an answer (e.g., normal temperature). Immediately after the start of the study, it became evident that we needed to iterate the CRF with an option for providers to designate if they were confident or not confident in certain responses. This iteration granted the provider situational adaptability to use their own clinical acumen to make the best possible assessment and treatment plan. Analyses of the data dependent, or independent, of the ‘confidence’ designation revealed contrasting performance of individual questions. For example, the ‘confident/not confident’ option for components of the virtual respiratory exam reduced false positive findings by 116 instances but increased missed clinical findings by 4 instances. This represents a 10-fold reduction in the ratio of false positives to false negatives. This finding suggests that future clinical decision support tools (paper or digital) must acknowledge the value of the ‘human’ aspect of clinical history taking and avoid approaches that marginalize telemedicine provider expertise.

Congruence between medication prescriptions generated from the virtual and in-person exams was high. The treatment plans were generated from multiple clinical components and this likely allowed for some redundancy. The in-person exam was associated with increased antibiotic stewardship for select cases. For example, amoxicillin was more often removed than added to a treatment plan after the in-person exam.

A scaled telemedicine model in resource-limited settings will likely rely on a subset of cases requiring an in-person exam. Next steps are to iterate the guidelines, improve interviewing techniques and evaluate portability in disparate settings. We anticipate that these steps will lead to a durable, scalable, and evidence-based pediatric telemedicine guideline. For workflows that include medication delivery, the path to scalability will likely require the continued referral of severe cases to emergency care, an in-person exam for moderate cases, and medication delivery alone for mild cases.

### Study limitations

These findings should be viewed within the context of the study limitations. First, this prospective cohort study was nested within a larger study to determine the feasibility and safety of the TMDS model^18^. The approach allowed for adaptive and iterative components based on unexpected logistical and clinical challenges; minor modifications were made to call in-take logistics, and clinical approaches and resources. Second, call center providers improved their interviewing skills over time as their familiarity with the clinical decision support tools increased. This may have impacted clinical guideline performance over time. Third, provider in-person exams were not performed for cases virtually categorized as severe to avoid delaying care. The ‘true’ status of these cases was unknown, therefore, the performance of the clinical tools in these situations could not be evaluated. Fourth, the low sample sizes for several clinical scenarios resulted in limited inference ability and/or wide confidence intervals.

## CONCLUSIONS

This study, and resulting guideline, represent a formative step towards an evidence-based pediatric telemedicine guideline for resource-limited settings. In-person exams for select cases remain important to address limitations with virtual exams and identify cases for escalation. Empowering providers to score their confidence in a response allowed for essential situational adaptability.

## Supporting information

S1 Clinical Guidelines

S2 Case Report Form

S3 Medication Formulary

STable 1

## Data Availability

All data produced in the present study are available upon reasonable request to the authors.

## Acknowledgements

We are grateful for the participation of the patients and their parents/guardians, as well as the field team who collected the data for this study. We would like to thank Randy Autrey and Krista Berquist for their administrative support, Glenn Morris at the Emerging Pathogens Institute and Desmond Schatz interim Chair of the Department of Pediatrics at the University of Florida for their academic support, as well as the Ministry of Public Health and Population (Ministère de la Santé Publique et de la Population - MSPP) for their partnership.

## Co-author contact information

Molly B. Klarman, MPH

Departments of Pediatrics and Environmental and Global Health, University of Florida,

Gainesville, FL, USA

Xiaofei Chi, MS

Department of Pediatrics, University of Florida, Gainesville, FL, USA

Youseline Cajusma

Departments of Pediatrics and Environmental and Global Health, University of Florida,

Gainesville, FL, USA

Katelyn E. Flaherty

Departments of Emergency Medicine and Environmental and Global Health,

University of Florida, Gainesville, FL, USA

Anne Carine Capois, M.D.

Departments of Pediatrics and Environmental and Global Health, University of Florida,

Gainesville, FL, USA

Michel Daryl Vladimir Dofiné, M.D.

Departments of Pediatrics and Environmental and Global Health, University of Florida,

Gainesville, FL, USA

Lerby Exantus, M.D.

Université d’État d’Haiti-Faculté de Médecine et de Pharmacie, Port-au-Prince, Haiti

Jason Friesen, MPH

Trek Medics International, Charlotte, NC, USA

Valery M. Beau de Rochars, M.D., MPH

Department of Health Services Research, Management and Policy, College of Public Health and Health Professions, University of Florida, Gainesville, FL, US

Torben K. Becker, M.D., Ph.D.

Departments of Emergency Medicine and Environmental and Global Health,

University of Florida, Gainesville, FL, USA

Chantale Baril, M.D.

Université d’État d’Haiti-Faculté de Médecine et de Pharmacie, Port-au-Prince, Haiti

Matthew J. Gurka, Ph.D.

Department of Pediatrics, University of Florida, Gainesville, FL, USA

Eric J. Nelson, M.D., Ph.D.

Departments of Pediatrics and Environmental and Global Health, University of Florida

Gainesville, FL, USA

