## Supplementary material for "Development and Evaluation of a Clinical Guideline for a Pediatric Telemedicine and Medication Delivery Service: A Prospective Cohort Study in Haiti": S1 Clinical Guidelines

**Supplementary Text 1.** Clinical Guidelines

These guidelines were written to be used within the scope of this pilot only

**A. WEIGHT ESTIMATES**

Most pediatric treatment plans require weight-based dosing. There are three equations to estimate the weight:

- For 0-12 months (new APLS): weight in kg = [ age in months x 0.5 ] + 4.
- For 1-5 years (old APLS formula): weight in kg = [ age in years + 4 ] x 2.
- For 6-12 years (Luscombe and Owen Formula): weight in kg = [ age in years x 3 ] + 7.

**Decision support.** Decision support tools will be provided to the medical team. These will include the WHO IMCI guidelines, American Academy of Pediatrics, Pediatric Telephone Protocols (Schmitt, 16th Edition), pediStat (calculates medication doses), and the Outbreak Responder rehydration calculator developed by Dr. Nelson et al.

**B. CLINICAL MANAGEMENT BY PROBLEM**

**#1. Fever with or without a source.**

The World Health Organization (WHO) IMCI guidelines divide management of fever between children less than two months of age and patients two months and older. The reason is that young infants can be at risk of life-threatening illness and may show very subtle symptoms and signs of impending death. Therefore, escalation of care is often recommended for infants less than 2 months of age. Most call reports of fever by parents on the phone will be subjective (not measured). These subjective fevers will be considered a legitimate fever until proven otherwise.

The phrase 'without' source refers to patients that do not have a clear explanation for their fever (e.g., viral cold with nasal discharge, pain from urination from a bacterial urinary tract infection, skin infection, invasive diarrhea with blood in stool). This section prioritizes care for patients with fever/low temperature (hypothermia) without a source. For those patients 2 months and older with a source, the recommendations for that specific problem are listed in other sections. Be mindful that young children with urinary tract infections will present with fever without a source because they will not complain, or be able to complain, of pain with urination. If this is a concern, they need to be seen at a clinic for a urine test in the morning.

**Less than 2 months of age.**

1. **RED. Danger sign or general clinical concern** (not feeding well, convulsions, fast breathing of 60 breaths per minute or faster), severe chest indrawing (retractions), measured fever (≥38 C rectal; ≥37.5 C oral), measured low temperature (less than 35.5 rectal or oral), movement only when stimulated (lethargic) or no movement (unresponsive). The case report form provides age-specific parameters for high and low rates; see vital sign table.

   Action. Will need immediate referral to hospital.

   Follow-up. Not applicable (will be at hospital).

2. **YELLOW / GREEN. No danger sign and no general clinical concern.**

   Action: Given that danger signs include a fever, most cases will be referred to a hospital. If there is borderline fever and mild family/clinical concern, these cases can be monitored, supported (paracetamol and ORS) and seen at the clinic in the morning. Give 1 liter of ORS via bottle to bottle fed patients as needed to avoid dehydration; breastfed patients should continue to breastfeed. The patient should take the ORS only if they are unable to feed normally. Once made, keep for only 8 hours.

   Medication/fluid dosing.

   Paracetamol: 15 mg/kg per dose by mouth every 6 hours as needed for fever and discomfort.

   ORS: Use as needed to avoid dehydration.

   Follow-up. Have the patient follow up with the clinic in the morning.

**Older than 2 months of age.**

1. **RED. Danger sign or stiff neck.** See above and case report form for more information.

   Action. Will need immediate referral to hospital.

   Follow up. Not applicable (will be at hospital).

2. **YELLOW / GREEN. No danger sign and no stiff neck.** See above and case report form for more information.

Action.

Give paracetamol for fever.

Give 1 liter of ORS to all patients to be used as needed to avoid dehydration. The patient should take the ORS only if they are unable to feed normally. Once made, keep for only 8 hours.

If there is a source, see that specific section for antibiotic recommendations.

Medication/fluid dosing.

Paracetamol: 15 mg/kg per dose by mouth every 6 hours as needed for fever and discomfort.

ORS: Use as needed to avoid dehydration.

Follow-up: If there is no improvement by the morning, have the patient follow up with the clinic that morning.

**#2. Breathing problem / Cough.**

The WHO IMCI guidelines focus on identifying if the patient has respiratory distress or respiratory failure. These are assessed by looking for signs of work of breathing: fast breathing, chest indrawing showing ribs (retractions), abnormal sound when the child breaths in (e.g., stridor), wheezing when the child breaths out (e.g., asthma), nasal flaring or head bobbing. Causes of breathing problems are bacterial pneumonia, asthma attack, stridor from croup (upper airway inflammation), foreign body (object in airway), and common viral respiratory infection/cold.

1. **RED. Danger sign, when patient is calm and has abnormal sounds when breathing in (stridor), severe fast breathing (under 1 year ≥60 breaths per min; 1 year or older ≥50 breaths per min) and/ or has three signs of distress (head bobbing, chest indrawing, nasal flaring). The case report form provides age-specific parameters for high and low respiration rates; see also vital sign table.**

   Action. Will need immediate referral to hospital.

   Follow up: Not applicable (will be at hospital)

2. **YELLOW. No danger sign but mild fast breathing for age and/or two signs of distress (head bobbing, chest indrawing, or nasal flaring). Upper limit for mild fast breathing: under 1 year 50 to less than 60 breaths per min; 1 year or older <40 to <50 breaths per min.**

   Action.

   If fever (measured either at home or by nurse) and cough, give amoxicillin.

   If 3 or more years old with wheeze or asthma history, give salbutamol.

   If less than 3 years old with wheeze or asthma history, patient goes to hospital because salbutamol needs a nebulizer.

   If less than 1 year old and significant mucus, suction the nose with a bulb and saline drops.

   Give 1 liter of ORS to all patients to be used as needed to avoid dehydration. The patient should take the ORS only if they are unable to feed normally. Once made, keep for only 8 hours.

   Provide supportive care with safe remedies for sore throat or cough.

   Medication/fluid dosing.

   Amoxicillin: 40 mg/kg per dose by mouth twice daily for 5 days. Maximum single dose is 1000 mg.

   Salbutamol: (100 ug/puff) two puffs with a spacer every 4 hours for 5 days.

   Paracetamol: 15 mg/kg per dose by mouth every 6 hours as needed for fever and discomfort.

   ORS: Use as needed to avoid dehydration.

   Follow-up: If there is no improvement by the morning, have the patient follow up with the clinic that morning. Patients over 3 years of age with wheeze, should follow-up with the clinic in the morning.

3. **GREEN. No danger sign.**

   Action:

   If fever (measured either at home or by nurse) and cough, give amoxicillin.

   If 3 or more years old with wheeze or asthma history, give salbutamol.

   If under 3 years old, needs morning clinic follow-up.

   If less than 1 year old and significant mucus, suction the nose with a bulb and saline drops.

   Provide supportive care with safe remedies for sore throat or cough.

Give 1 liter of ORS to all patients to be used as needed to avoid dehydration. The patient should take the ORS only if they are unable to feed normally. Once made, keep for only 8 hours.

Provide supportive care with safe remedies for sore throat or cough.

Medication/fluid dosing

Amoxicillin: 40 mg/kg per dose by mouth twice daily for 5 days. Maximum single dose is 1000 mg.

Salbutamol: (100 ug/puff) two puffs with a spacer every 4 hours for 5 days.

Paracetamol: 15 mg/kg per dose by mouth every 6 hours as needed for fever and discomfort.

ORS: Use as needed to avoid dehydration.

Follow-up: Follow up if patient remains without danger signs and does not worsen. For patients under 3 years of age with wheeze, morning follow-up is required.

**#3. Dehydration / Vomit (3A) / Diarrhea (3B).**

The WHO IMCI guidelines address the assessment and treatment of dehydration in the context of diarrheal disease. However, methods of rehydration for diarrheal disease can also be used to address dehydration from other causes, including vomiting. Determining if a child has tears (best for children under 5 years) or has urinated (all ages) in the last 8 hours can be useful to determine risk of dehydration. With respect to vomiting, vomiting alone is not dangerous, especially if it follows a cough in a young child. Dark green vomit however can be a sign of intestinal obstruction. Multiple episodes in 24 hours (approximately above six), and/or if child cannot take liquids, can lead to dehydration and electrolyte problems.

Assessment. The assessment of dehydration is based on four clinical features: general condition (well/alert, restless/irritable (<5 years only), lethargic/unconscious), eyes (normal/sunken), thirst (normal, drinks eagerly/thirsty, not able to drink/drinks poorly), skin turgor (goes back quickly in less than two seconds, slowly in two to three seconds, very slowly greater than three seconds). These signs are scored ‘No’, ‘Some’, and ‘Severe’ to approximate 0-4%, 5-9%, and ≥10% weight loss by scoring two features in the highest category.

SEVERE (≥10). At least two signs of Severe dehydration (10+%): Lethargy/unconscious, eyes very sunken, drinks poorly/ unable to drink and/or skin pinch goes back very slowly/ >3 seconds. This is RED.

SOME (5-10%). At least two signs of Some dehydration (5-10%): Restless/irritable (only for < 5 years), sunken eyes, drinks eagerly/ thirsty, and/ or skin pinch goes back slowly/ 2-3 seconds. This is YELLOW.

NO (0-5%): Default if above criteria are not met. This is GREEN.

Treatment:

1. **RED. Danger sign, severe dehydration and/or dark green vomit**.

   Action. Will need immediate referral to hospital.

   Follow up. Not applicable (will be at hospital)

2. **YELLOW. No danger sign and Some dehydration.**

   Action

   Give ORS. If patient has vomiting. Give more frequent ORS in smaller volumes.

   Give antibiotics if indicated (read below).

   Consider anti-vomiting medication if available and vomiting is severe.

   Medication/fluid dosing.

   Give ORS. The volumes for correction are as follows:

   - Volume (<1 yr): Correct with 75 ml/kg over 6 hours.
   - Volume (≥1 yr): Correct with 75 ml/kg over 4 hours.
   - Above 70 kg, the calculations are set to 70 kg (approximately 5 liters)

     While correcting for dehydration, ongoing losses must be given to replace the equivalent volume lost: Less than 2 years, 50-100 ml of ORS after each loose stool. Greater than 2 years, 100-200 ml of ORS after each loose stool.

   Give Antibiotic for patients with acute watery diarrhea that looks like rice-water and is concerning for cholera (no blood).

Below 8 years: Erythromycin 12.5 mg/kg per dose by mouth four times a day for 3 days; maximum single dose is 500 mg. Note that Doxycycline can alternatively be given to children less than 8 years per WHO guidelines.

Above 8 years: Doxycycline 4 mg/kg mg by mouth once; maximum dose is 300 mg.

Give azithromycin for patients with stool/diarrhea with blood.

Azithromycin: 10 mg/kg by mouth once a day for 5 days; maximum single dose 500 mg.

Alternative: Ciprofloxacin per dose 15 mg/kg by mouth twice a day for three days; maximum single dose is 500 mg.

Give zinc. For 2 months to <6 months, 10 mg by mouth once a day for 10 days. For 6 months to five years, 20 mg by mouth once a day for 10 days.

Give ondansetron (if available and for severe emesis or nausea). For 1 year to <4 years, 2 mg by mouth once. For 4 years and above, 4 mg by mouth once.

Follow-up. If there is no improvement by the morning, have the patient follow up with the clinic that morning.

3. **GREEN. No danger sign and No dehydration.**

Action.

Give ORS. If patient has vomiting, give more frequent ORS in smaller volumes. For children that don't have diarrhea but do not have tears or have not urinated in the last 8 hours, give 1-2 packets of ORS.

Give antibiotics only for patients with blood in the stool (not for acute watery diarrhea).

Consider anti-vomiting medication if available and vomiting is severe.

Medication/fluid dosing.

Give ORS. Ongoing losses from diarrhea are replaced with the equivalent volume lost. Similarly, each episode of emesis can be replaced with an equal volume of ORS but space in smaller volumes over time.

- Less than 2 years, 50-100 ml of ORS after each loose stool.
- Greater than 2 years, 100-200 ml of ORS after each loose stool.

Give azithromycin for patients with stool/diarrhea with blood.

Azithromycin: 10 mg/kg by mouth once a day for 5 days; maximum single dose 500 mg.

Alternative: Ciprofloxacin 15 mg/kg per dose by mouth twice a day for three days; maximum single dose is 500 mg.

Give ondansetron (if available and for severe emesis or nausea): For 1 year to 4 years, 2 mg by mouth once. For 4 years and above, 4 mg by mouth once.

Follow-up: No need for follow up if patient remains without danger signs and does not worsen.

**#4. Ear pain.**

Ear pain in pediatrics is typically caused by fluid congestion in the inner ear and/or inflammation from a viral infection, and less often from a bacterial infection. It is typically not an emergency unless there is infection into the surrounding tissues. Ear pain can also be referred from pain in the mouth.

1. **RED.** Pain when you press the bone behind the ear may indicate mastoiditis that will require hospital level care. The area may be red, warm, and swollen in mastoiditis (infection of the mastoid boney space).
2. **YELLOW.** Acute infection: Ear pain and/or puss draining from the ear for less than 14 days. Give amoxicillin 40 mg/kg per dose by mouth twice daily for 5 days. Treat pain with acetaminophen and/or ibuprofen.

   Chronic Infection: These should be managed at a clinic. Puss is draining for more than 14 days. Place cotton wick in the ear to drain the puss. Use topical quinolone drops for 14 days.
3. **GREEN.** If there is no ear pain and no puss, then no treatment is needed.

   Follow-up: Yellow follow-up at clinic if no improvement. Green follow-up as needed.

**#5 Skin problem.**

Skin problems may be caused by an allergy after an exposure (e.g., a specific food with a known food allergy), infection (e.g., fungus, virus, bacteria) or irritation from an insect (e.g., scabies, bed bugs). Most of these problems will not require seeking care at night. However, rapidly spreading infections or infections near the mouth, eyes, or genitalia may require attention. A bacterial infection is characterized by being red, warm, raised, and painful; not all infections are with fever; they are also often a specific location with or without puss. The size of the infection is important (coin-size, size of a hand, larger than the size of a hand). Sometimes fluid forms below the skin causing an abscess that might be infected with bacteria.

1. **RED**. Skin infections that are spreading quickly and/or involve the eye, mouth, and genitalia (other than mild fungal diaper rash) may need immediate care at the hospital. Large infections (larger than the size of the hand) that might be bacterial may need hospital level care. Allergic reactions that have abdominal pain, swollen tongue, severe hives, or lips require immediate loratadine and/or diphenhydramine and transfer to the hospital for concern of anaphylaxis.
2. **YELLOW**. Skin infections that are specific to one part of the body and are red, warm, raised, and painful with fever are likely to be bacterial. Initiate treatment with cephalexin (25 mg/kg per dose orally twice a day for five days). Alternatively, a secondary choice is co-trimoxazole (trimethoprim 4 mg/kg per dose by mouth every 12 hours; dosed by trimethoprim component) for 5 days. Allergic reactions with moderate hives but no abdominal pain, swollen tongue or lips can be treated with loratadine or diphenhydramine.
3. **GREEN**. All other cases do not necessitate night-time evaluation and should be followed up at a daytime clinic. If a non-urgent skin problem is found at the household visit, medications available are permethrin (scabies), miconazole (fungal diaper rash), and hydrocortisone (minor inflammation). Diphenhydramine can help to address itchy skin, but it lasts for only a few hours, does not solve the problem, and makes patients sleepy. Allergic reactions with mild hives but no abdominal pain, swollen tongue or lips can be treated with loratadine. Lice and Scabies can be treated with permethrin or benzyl benzoate. For infants, dilute 15% benzyl benzoate one-part to one-part in water, or dilute 25% benzyl benzoate one-part to three-parts in water. Severe scabies infections with pustules can be treated with oral ivermectin only for patients that weigh at least 15 kg (0.2mg/kg per dose by mouth once; repeat in two weeks).

   Follow-up: Yellow follow-up at clinic if no improvement. Green follow-up as needed.

**#6 Urinary tract infection.**

Older patients may have pain with urination (dysuria). However, young patients have are likely to have nonspecific signs. Symptoms may include fever without a source, vomiting and poor feeding, lethargy, irritability, abdominal pain, and pain on side of the lower back. Infants with fever under two months of age will be referred to the hospital based on the fever guidelines.

1. **RED**. Danger signs. Immediately seek care at the hospital.
2. **YELLOW**. For pain with urination and/or lower back pain on the side. Give oral co-trimoxazole (trimethoprim 10 mg/kg by mouth every 12 hours; dosed by trimethoprim component) for 5 days. Alternatively, cephalexin can be given (25 mg/kg per dose by mouth every 6 hours for 5 days; maximum dose 2000 mg). If there is streptococcus pharyngitis and a urinary tract infection, consider amoxicillin/clavulanic acid 20 mg/kg by mouth every 8 hours for 10 days (dosing based on amoxicillin component).
3. **GREEN**. For pain with urination. Give oral co-trimoxazole (trimethoprim 10 mg/kg by mouth every 12 hours; dosed by trimethoprim component) for 5 days. Alternatively, cephalexin can be given (25 mg/kg per dose by mouth every 6 hours for 5 days; maximum dose is 2000 mg). If there is streptococcus pharyngitis and a urinary tract infection, consider amoxicillin/clavulanic acid 20 mg/kg by mouth every 8 hours for 10 days (dosing based on amoxicillin component).

   Follow-up: Yellow follow-up at clinic if no improvement. Green follow-up as needed.

**E. Other diagnosis.** The management of other less common diagnoses will not be scripted. The approach will rely on standard of care by nurses, scope of practice for nurses, and consult from the on-call doctor. The intentionally short list of available medications will also constrain the scope of management to pediatric urgent care needs. One example is an allergic reaction. Using the same triage approach (RED, YELLOW, GREEN), triage the situation. If there is allergic reaction with involvement of the mouth or ability to breath, the patient is RED and needs to go to the hospital immediately; dose with diphenhydramine and loratadine during transport. If there is a full body allergic reaction without involvement of the mouth or breathing the patient is YELLOW and diphenhydramine and loratadine can both be used. Follow-up with clinic in the morning if no improvement. Be aware these medications will make the patient tired. If there is an allergic reaction on a specific body part without involvement of the mouth or breathing the patient is GREEN and diphenhydramine or loratadine can both be used. Be aware diphenhydramine will make the patient tired. A second example is Strep throat; symptoms are typically fever, swollen lymph nodes along the side of the neck, no cough, and white painful lesions on the back of the throat. The dose of amoxicillin is the same (40 mg/kg per dose twice a day) but the course is 10 days. For each of these situations use a similar method for follow-up: Yellow follow-up at clinic if no improvement. Green follow-up as needed.

**Appendix**

**Normal Vital signs***

| Age | 1-7 days | 1-3 weeks | 1 month | 6 months | 12 months | 18 months |
|---|---|---|---|---|---|---|
| Weight | 3 kg | 3 kg | 4 kg | 7 kg | 10 kg | 11 kg |
| Heart Rate | 90-165 | 105-180 | 120-180 | 100-180 | 100-180 | 100-180 |
| Resp. Rate | 30-60 | 30-60 | 30-60 | 30-60 | 24-40 | 24-40 |

| Age | 2 years | 3 years | 5 years | 6 years | 8 years | 10 years |
|---|---|---|---|---|---|---|
| Weight | 12 kg | 14 kg | 18 kg | 25 kg | 31 kg | 37 kg |
| Heart Rate | 60-140 | 60-140 | 60-140 | 60-140 | 60-140 | 60-140 |
| Resp. Rate | 24-40 | 24-40 | 22-34 | 18-30 | 18-30 | 18-30 |

*Fever: oral >37.5 C; rectal >38 C. Oxygen: Normal ≥90%. Normal values derived from PALS and Stanford (LPCH) Pediatric Emergency Guidelines. Weight estimates explained at the top of the guidelines. WHO IMCI thresholds set fast breathing at 50 breaths per minute (<1 year) and 40 breaths per minute (≥1 year).
