## Supplementary material for "Development and Evaluation of a Clinical Guideline for a Pediatric Telemedicine and Medication Delivery Service: A Prospective Cohort Study in Haiti": S2 Case Report Form

Supplementary Text 2. Case Report Form

**INACT2 CASE REPORT FORM/ DECISION SUPPORT TOOL** **Patient ID __ -__ __ __ __ -__** ['P/X'-####A]

### IDENTIFICATION AND LOGISTICS

**IDENTIFICATION** **NURSE ID ___Initials _______**

| | | | |
|---|---|---|---|
| 1. __ __/ __ __/__ __<br>2C. Duration call<br>___ ___ min | 2A. Time: _ _ : _ _ am ☐ pm☐<br>2B. Repeat caller: ☐ No ☐ Yes<br>dd _ _/ mm _ _ | 3. < 2 years: __ __ months<br>≥ 2 years: __ __ years | 4. Location |
| 5. Caller Name (First/ Last) | | | 6. Primary Mobile<br>509 _ _ _ _ -_ _ _ _ |
| 7. Patient Name (First/ Last) | | 8. Sex: ☐ F ☐ M | 9. Secondary Mobile<br>509 _ _ _ _ -_ _ _ _ |
| 10A. Problem(s)<br>10B. Duration:<br>__ __ days | ☐ Fever #1 ☐ Cough/ Breathing #2 ☐ Vomit #3A<br>☐ Diarrhea #3B ☐ Ear Pain #4 ☐ Skin #5<br>☐ Pain with urine #6 ☐ Other #7: (free text) | | 11. Landmarks |

### 24-hour Follow Up

☐ Need ☐ Not Needed

**Required for RED patients and those referred to a clinic** **NURSE ID ___Initials _______**

| | | | |
|---|---|---|---|
| First call attempt | Time: _ _ : _ _ am ☐ pm☐ | dd _ _mm _ _yy _ _ | ☐ not completed ☐ completed |
| Second call attempt | Time: _ _ : _ _ am ☐ pm☐ | dd _ _mm _ _yy _ _ | ☐ not completed ☐ completed |
| 12. Condition | Current status: ☐ Well ☐ Sick/Improving ☐ Sick/Same ☐ Sick/Worsening ☐ Died ☐ Unknown | | |
| 13A. Care sought | Level of care patient first sought | | ☐ Hospital ☐ Clinic ☐ None ☐ Other:_ |
| 13B. Problem | If yes, what did they consider the main problem? | | ☐ Same ____(#) ☐ Different _______ |
| 13C. Intervention | If care sought, what was done? | | |
| 14. Patient location | Is the patient still receiving care? Where? | | ☐No (Home) Yes:☐ Hospital ☐ Other:_ |
| 15. Comments | | | |

### 10-day Follow Up

**Required for all patients** **NURSE ID ___Initials _______**

| | | | |
|---|---|---|---|
| First call attempt | Time: _ _ : _ _ am ☐ pm ☐ | dd _ _mm _ _yy _ _ | ☐ not completed ☐ completed |
| Second call attempt | Time: _ _ : _ _ am ☐ pm ☐ | dd _ _mm _ _yy _ _ | ☐ not completed ☐ completed |
| Third call attempt | Time: _ _ : _ _ am ☐ pm ☐ | dd _ _mm _ _yy _ _ | ☐ not completed ☐ completed |
| 16. Condition | Current status: ☐ Well ☐ Sick/Improving ☐ Sick/Same ☐ Sick/Worsening ☐ Died ☐ Unknown | | |
| 17A. Care sought (1) | Level of care patient first sought | | ☐ Same as at 24 hour follow-up<br>☐ Hospital ☐ Clinic ☐ None ☐ Other:_ |
| 17B. Problem | If yes, what did they consider the main problem? | | ☐ Same ____(#) ☐ Different _______ |
| 17C. Intervention | If care sought, what was done? | | |
| 18A. Care Sought (2) | Was care sought a second time? | | ☐ Hospital ☐ Clinic ☐ None ☐ Other:_ |
| 18B. Problem | If yes, what did they consider the main problem? | | ☐ Same ____(#) ☐ Different _______ |
| 18C. Intervention | If care sought, what was done? | | |
| 19. Patient location | Is the patient still receiving care? Where? | | ☐ No (Home) Yes:☐ Hospital ☐ Other:_ |
| 20. Medication | Was medication prescribed taken? ☐ Yes ☐ None prescribed ☐ No, why? | | |
| 21. Comments | | | |

**Feedback required for all patients**

| | | |
|---|---|---|
| 22. Price of service appropriate (only those that receive delivery) ☐ NA ☐ Yes ☐ Too expensive | | |
| 23. Overall impression of delivery service | | ☐ NA ☐ Great ☐ Good ☐ Okay ☐ Poor ☐ Bad |
| 24. Overall impression of call service | | ☐ Great ☐ Good ☐ Okay ☐ Poor ☐ Bad |
| 25. Would call again? | ☐ Yes ☐ No<br>If no, why? | 26. Additional feedback: |

IRB Project #: myIRB 201802920
Version: 9/22/2020

### CALL CENTER TRIAGE

**NURSE ID___ Initials _______**

**DANGER SIGNS** (danger sign **stop** and send to the hospital. Plan A. RED)

| | | |
|---|---|---|
| 27. General | Unresponsive (with stimulation) / lethargic | ☐ **YES** ☐ No |
| 28. Hydration | Able to drink / breastfeed<br>(Effectively drank / breastfed without major vomiting) | ☐ **NO** ☐ Yes |
| 29. Neurologic | Has/does the child have a seizure (last 24 hours). | ☐ **YES** ☐ No |

**VITAL SIGNS (see vital sign table for reference)**

| | | |
|---|---|---|
| 30. Temp / fever | Under 1 year measured rectally (fever ≥38C / 100.4F)<br><br>Above 1 year orally (Fever ≥ 37.5C / 99.5F; uncorrected) | Fever: ☐YES (Subj), ☐ No (subj)<br>☐ Yes (Obj), ☐ No (obj)<br>Measurement: _ _ _ . _ ☐ C ☐ F<br>☐ Oral ☐ Axillary ☐ Rectal |
| 31A.Respiration Rate | Count chest rises in 15 seconds<br>• Under 1 year **>= 60** breaths/min, hospital<br>• 1 year or older **>=50**, hospital<br>• If < 2 yr and **<24** breaths/min, hospital<br>• If >= 2 yr and **<18** breaths/min, hospital | _ _ per 15 second x 4 for rpm<br>_ _ breaths/min<br>☐ Not obtained/ unreliable |
| 31B. Fast breathing | Breaths per minute:<br>Under 1 year >= 50,<br>1 year or older >= 40<br>Fast breathing could be a sign of pneumonia | ☐ Yes ☐ No<br>☐ Not obtained/ unreliable |
| 32. Heart Rate | Only for children less than 5 years, parent counts for 15 seconds with hand over left chest<br>• If **>= 180** beats/min, hospital (all ages)<br>• If < 2 yr and **<100** beats/minute, hospital<br>• If >= 2 yr and **<60** beats/minute, hospital | _ _ _ per 15 second X 4 for bpm<br>_ _ _ beats/min<br>☐ Not obtained/unreliable |

### CALL CENTER PROBLEM SPECIFIC QUESTIONS

| **Breathing problem / Cough #2.** | | | | | **CONFIDENT** |
|---|---|---|---|---|---|
| 33. Head | Head goes up and down when breathing more than normal ('head bobbing') | ☐ YES | ☐ No | ☐ Unknown | ☐ Yes ☐ No |
| 34. Nose | Do nostrils go in and out when breathing more than normal ('nasal flaring') | ☐ YES | ☐ No | ☐ Unknown | ☐ Yes ☐ No |
| 35. Nose | Nasal discharge. Mucus is a sign of infection (a source) | ☐ YES | ☐ No | ☐ Unknown | NA |
| 36. Mouth | Is there a cough | ☐ YES | ☐ No | ☐ Unknown | NA |
| 37. Neck/Chest: | Chest indrawing showing ribs when breathing (retractions) more than normal | ☐ YES | ☐ No | ☐ Unknown | ☐ Yes ☐ No |
| 38. Neck: Stridor | When calm, abnormal sound when breaths in (Stridor) that is different than the normal sounds | ☐ **YES** | ☐ No | ☐ Unknown | ☐ Yes ☐ No |
| 39. Chest: Wheeze | When calm, wheeze when child breaths out (may be Asthma) that is different than the normal or nasal mucus sounds | ☐ YES | ☐ No | ☐ Unknown | ☐ Yes ☐ No |
| If confident in # 33, 34, and 37, treat at hospital as red if all three are yellow. If confident in #38, treat at hospital. | | | | | |

### CONTINUED CALL CENTER QUESTIONS

**DEHYDRATION (all patients)**

| | | |
|---|---|---|
| 40A. Urine (all ages) | Was there urine in the last 8 hours? | ☐ Yes ☐ No ☐ Unknown |
| 40B. Tears (<5 years) | Tears present? (with or without crying) | ☐ Yes ☐ No (do 42-46) ☐ Unknown |
| 41A. Vomiting **(#3A)** | Episodes in last 24 hours: ☐ None ☐ 1-2 ☐ 3-5 ☐ 6-11 ☐ 12 or more ☐ Unknown | |
| 41B. Vomiting | Is vomit dark green vomit, If yes **Red**. | ☐ Yes ☐ No ☐ Unknown |

**DEHYDRATION (diarrhea patients) #3B** **CONFIDENT**

| | | | | |
|---|---|---|---|---|
| 42A. General: ≥ 5 y | ☐ Well/Alert | --------BLANK-------- | ☐ Lethargic/ Unconscious **Red** | ☐ Yes ☐ No |
| 42B. General: < 5 y | ☐ Well/Alert | ☐ Restless/irritable | ☐ Lethargic/Unconscious **Red** | ☐ Yes ☐ No |
| 43. Eyes | ☐ Not Sunken | --------BLANK-------- | ☐ Sunken | ☐ Yes ☐ No |
| 44. Thirst | ☐ Normal | ☐ Drinks eagerly/thirsty | ☐ Not able to drink (effectively) | ☐ Yes ☐ No |
| 45. Skin pinch | ☐ Normal (<2 sec) | ☐ Slow (2-3 sec) | ☐ Very slow (>3 sec) | ☐ Yes ☐ No |
| 46. Assess by scoring highest 2 findings | ☐ Green (<5%; No)<br>Score only those items marked as "Yes" for confident in the answer. | ☐ Yellow (5-9%; Some) | ☐ **Red** (≥10%; Severe, hospita | |

| | | | | |
|---|---|---|---|---|
| 47. Loose stools | Loose stools in last 24 hours: ☐ 1-2 ☐ 3-5 ☐ 6-11 ☐ 12 or more | | | |
| 48. Rice-water stool | Is it like rice-water (see plan for antibiotics). | ☐ Yes | ☐ No | ☐ Unknown |
| 49. Bloody stool | If yes, will require an antibiotic. | ☐ Yes | ☐ No | ☐ Unknown |

**EAR PAIN #4**

| | | | | |
|---|---|---|---|---|
| 50. Puss from ear | If yes, antibiotics. If not, supportive care. | ☐ Yes | ☐ No | ☐ Unknown |
| 51. Pain behind ear | Is the bone behind the ear painful when pressed, red, and swollen. Consider mastoiditis (red), typically follows an ear infection. | ☐ **Yes** | ☐ No | ☐ Unknown |

**SKIN PROBLEM #5**

| | |
|---|---|
| 52A. Type | ☐ Allergic (itchy with an 'exposure') ☐ Bacterial Infection (red, raised, warm, pain)<br>☐ Scabies (itchy small bumps) ☐ Other (e.g., viral rash): |
| 52B. If bacterial, size | ☐ Size of a coin ☐ Size of a hand ☐ Larger than a hand (hospital or morning follow-up) |

**PAIN WITH URINATION #6**

| | | | | |
|---|---|---|---|---|
| 53. Painful urination | If yes and there is a fever, suggests urine infection. | ☐ Yes | ☐ No | ☐ Unknown |

**OTHER PROBLEMS #7** Ask questions that target the specific problem and gauge severity.

| |
|---|
| 54. Other problem |

**PAST MEDICAL HISTORY**

| | |
|---|---|
| 55. Past problems | |
| 56A. Medications:<br><br>56B. When was the last dose given? | 57. Allergies: ☐ No, ☐ Unknown, Yes: |

### HOUSEHOLD TRIAGE. NURSE ID___ Initials _______ ☐ REFUSED/ OUT OF AREA/ FAILED

**DANGER SIGNS** (danger sign **stop** and send to the hospital. Plan A. RED)

| | | |
|---|---|---|
| 58. General | Unresponsive (with stimulation) / lethargic | ☐ **YES** ☐ No |
| 59. Hydration | Able to drink / breastfeed<br>(effectively drank/breastfed without major vomiting) | ☐ **NO** ☐ Yes |
| 60. Neurologic | Has/does the child have a seizure (last 24 hours) | ☐ **YES** ☐ No |

**VITAL SIGNS (see vital sign table for reference)**

| | | |
|---|---|---|
| 61. Temp / fever | Under 1 year measured rectally (fever ≥ 38 C)<br><br>Above 1 year orally (Fever ≥ 37.5C; uncorrected)<br><br>Do axillary temp on all (Fever ≥ 37.5C; uncorrected) | Fever: ☐ Yes (Obj), ☐ No (Obj)<br><br>☐ Oral ☐ Rect _ _ _ . _ ☐ C ☐ F<br>Axillary: _ _ _ . _ ☐ C ☐ F |
| 62A.Respiration Rate | Count chest rises in 15 seconds<br>• Under 1 year **≥60** breaths/min, hospital<br>• 1 year or older **≥50** breaths/min, hospital<br>• If < 2 yr and **<24** breaths/min, hospital<br>• If ≥ 2 yr and **<18** breaths/min, hospital | _ _ per 15 second x 4 for rpm<br>_ _ breaths/min ☐ Not obtained |
| 62B. Fast breathing | Breaths/min: Under 1 year >= 50, 1 year or older >= 40. Fast breathing could be a pneumonia sign. | ☐ YES ☐ No ☐ Unsure |
| 63. Heart Rate<br>(pulse-ox or palpation) | • If **>180** beats/min, hospital (all ages)<br>• If < 2 yr and **<100** beats/minute, hospital<br>• If ≥ 2 yr and **<60** beats/minute, hospital | _ _ _ per 15 second X 4 for bpm<br>_ _ _ beats/min ☐ Not obtained |

**VITAL SIGNS**

| | | |
|---|---|---|
| 64. Oxygen | If **<90%** while awake, **stop** and direct to hospital | _ _ _ % |
| 65. Weight | Measure with minimal clothing (as appropriate) | _ _ _ . _ _ kg |
| 66. MUAC | Malnutrit: 2mo-6 mo if **<110mm**; 6mo-<5 yr**<115mm** | _ _ _ mm |

### HOUSEHOLD PROBLEM SPECIFIC QUESTIONS

**Breathing problem / Cough #2.**

| | | |
|---|---|---|
| 67. Head | Does head go up and down when breathing more than normal ('head bobbing') | ☐ YES ☐ No ☐ Unknown |
| 68. Nose | Do nostrils go in and out when breathing more than normal ('nasal flaring) | ☐ YES ☐ No ☐ Unknown |
| 69. Nose | Nasal discharge. Mucus is a sign of infection (a source) | ☐ YES ☐ No ☐ Unknown |
| 70. Mouth | Is there a cough | ☐ YES ☐ No ☐ Unknown |
| 71. Neck/Chest: | Chest indrawing showing ribs when breathing (retractions) more than normal | ☐ YES ☐ No ☐ Unknown |
| 72. Neck: Stridor | When calm, abnormal sound when breaths in (Stridor) that is different from normal | ☐ **YES** ☐ No ☐ Unknown |
| 73. Chest: Wheeze | When calm, wheeze when breaths out (may be asthma, noisy noise) that is different than the normal sounds. | ☐ YES ☐ No ☐ Unknown |
| 74. Chest: Stethoscope | When listening with a stethoscope, abnormal breaths sounds? | ☐ Normal ☐ Wheeze (out)<br>☐ Crackles ☐ Other ☐ Unknown |
| | | If more than two yellow signs, treat at hospital as red. |

### CONTINUED HOUSEHOLD QUESTIONS

#### DEHYDRATION (all patients)

| | | | |
|---|---|---|---|
| 75A. Urine (all ages) | Was there urine in the last 8 hours? | ☐ Yes ☐ No | ☐ Unknown |
| 75B. Tears (<5 years) | Tears present? (with or without crying) | ☐ Yes ☐ No (do 77-81) | ☐ Unknown |
| 76A. Vomiting **(#3A)** | Episodes in last 24 hours: ☐ None ☐ 1-2 ☐ 3-5 ☐ 6-11 ☐ 12 or more ☐ Unknown | | |
| 76B. Vomiting | Is vomit dark green, If yes **Red** | ☐ Yes ☐ No | ☐ Unknown |

#### DEHYDRATION (diarrhea patients) #3B

| | | | |
|---|---|---|---|
| 77A. General: > 5 y | ☐ Well/Alert | --------BLANK-------- | ☐ Lethargic/ Unconscious **Red** |
| 77B. General: < 5 y | ☐ Well/Alert | ☐ Restless/irritable | ☐ Lethargic/ Unconscious **Red** |
| 78. Eyes | ☐ Not Sunken | --------BLANK-------- | ☐ Sunken |
| 79. Thirst | ☐ Normal | ☐ Drinks eagerly/thirsty | ☐ Not able to drink (effectively) |
| 80. Skin pinch | ☐ Normal (<2sec) | ☐ Slow (2-3 sec) | ☐ Very slow (>3 sec) |
| 81.Assess by scoring highest 2 findings | ☐ Green (<5%; No) | ☐ Yellow (5-9%;Some) | ☐ **Red** (≥10%; Severe, send to hospital) |

| | | | | | |
|---|---|---|---|---|---|
| 82. Loose stool | How many stools in the last 24 hours | ☐ 1-2 | ☐ 3-5 | ☐ 6-11 | ☐ 12+ |
| 83. Rice-water stool | Is it like rice-water (see plan for antibiotics) | ☐ Yes | ☐ No | ☐ Unknown | |
| 84. Bloody stool | If yes, will require an antibiotic | ☐ Yes | ☐ No | ☐ Unknown | |

#### EAR PAIN #4

| | | | | |
|---|---|---|---|---|
| 85. Puss from ear | If yes, antibiotics. If not, supportive care | ☐ Yes | ☐ No | ☐ Unknown |
| 86. Pain behind ear | Is the bone behind the ear painful when pressed, red, and swollen. This could be mastoiditis (red). | ☐ **Yes** | ☐ No | ☐ Unknown |

#### SKIN PROBLEM #5

| | |
|---|---|
| 87A. Type | ☐ Allergic (itchy with an 'exposure') ☐ Bacterial Infection (red, raised, warm, pain)<br>☐ Scabies (itchy small bumps) ☐ Other (e.g., viral rash): |
| 87B. If bacterial, size | ☐ Size of a coin ☐ Size of a hand ☐ Larger than a hand (hospital or morning follow-up) |

#### PAIN WITH URINATION #6

| | | | | |
|---|---|---|---|---|
| 88. Painful urination | If yes and there is a fever, suggests urine infection. | ☐ Yes | ☐ No | ☐ Unknown |

#### OTHER PROBLEMS #7 Ask questions that target the specific problem and gauge severity.

| |
|---|
| 89. Other problem |

#### PAST MEDICAL HISTORY

| | |
|---|---|
| 90. Past problems | |
| 91. Medications (current): | 92. Allergies: ☐ No, ☐ Unknown, Yes: |

### ASSESSMENT AND PLAN

#### CALL CENTER

| | | |
|---|---|---|
| 93. PLAN | ☐ **A. Mild** ☐ **B. Moderate** ☐ **C. Severe** (hospital) | |
| 94. Fever #1 (☐ not needed) | Paracetamol 15 mg/kg every 6 hours as needed for fever. | |
| 95. Breathing/ Cough#2 (☐ not needed)<br><br>Note: It is ok to refill salbutamol/ albuterol for known asthmatics that are now without medication. | Pneumonia (fever and cough): Amoxicillin 40 mg/kg per dose by mouth twice daily for 5 days | |
| | Asthma (wheeze): If < 3 years with wheeze, yellow goes to hospital and green needs morning clinic follow-up. If ≥ 3 years, 100 ug/puff 2 puffs with spacer every 4 h for 5 days for wheeze. | |
| | If <1 yr and significant mucus, provide bulb suction and saline drops. | |
| 96. Dehydration<br>Vomit #3A<br>Diarrhea #3B<br>(☐ not needed) | ORS Correction (75ml/kg), Amount = _ _ _ ☐ L ☐ Packets; ☐ over 4 hrs ☐ over 6 hrs<br>ORS Maintenance, Amount = _ _ _ ☐ Liters ☐ Packets. [½ glass to 1 glass per loose stool or vomit; also give ORS for patients with hydration concerns – no tears/urine] | |
| | Diarrhea. For cholera, antibiotics are for 'Some'/'Severe'. Don't delay transport for antibiotics.<br>Watery**:** <8 yrs: Erythromycin 12.5mg/kg per dose by mouth 4 x a day for 3 days; max dose is 500mg.<br>Watery**:** ≥8 yrs: Doxycycline 4 mg/kg mg by mouth once; max dose is 300 mg.<br>Bloody: (all ages). Azithromycin: 10 mg/kg by mouth once a day for 5 days; max dose is 500 mg. | |
| | Diarrhea present. Zinc**:**<br>☐ 2 mo-<6 mo. 10 mg by mouth once a day for 10 days.<br>☐ 6 mo-<5 yrs. 20 mg by mouth once a day for 10 days. | |
| 97. Ear Pain #4 (☐ not needed) | First choice (ear pain <14 days). Amoxicillin: 40 mg/kg per dose by mouth twice daily for 5 days. | |
| 98. Skin #5 (☐ not needed) | Allergic (Benadryl/hydrocortisone). Infection (cephalexin). Scabies (benzyl benzoate). | |
| 99. Pain with urine #6 (☐ not needed) | Co-trimoxazole: 10 mg/kg trimethoprim and 40 mg/kg sulfamethoxazole per dose twice a day for 5 days. If pneumonia is also present, use amoxicillin/clavulanic acid. | |
| 100. Other problem (#7) (☐ not needed) | Medications (see clinical guideline appendix). Note: Strep throat requires 10 days of amoxicillin. | |
| 101. Additional actions (☐ not needed) | | |
| 102. Where to treat? | ☐ Hospital ☐ Household (yellow/green) | 103. If in 5 km, delivery needed:<br>☐ Yes, ☐ No |
| 104. Morning follow-up | ☐ Clinic (mandatory)<br>☐ Clinic if not improved (most yellow)<br>☐ Clinic as needed (most green)<br>☐ Not applicable | 105. Notes |

#### CHANGES MADE AT THE HOUSEHOLD ☐ REFUSED/ OUT OF AREA/ FAILED

| | | |
|---|---|---|
| 106. RE-ASSESSMENT | ☐ No change ☐ **A. Mild** ☐ **B. Moderate** ☐ **C. Severe** (hospital). | |
| 107. Medication change | ☐ No change Problem type (____________________) and assessment change:<br>Medication change: | |
| 108. Where to treat? | ☐ No change ☐ Hospital<br>☐ Household (yellow/green) | 109. Time nurse returns to the office:<br>__ __ : __ __ am ☐ pm☐ |
| 110. Morning follow-up | ☐ No change<br>☐ Clinic (mandatory)<br>☐ Clinic if not improved (most yellow)<br>☐ Clinic as needed (most green) | 111. Notes |
| | | 112. GPS N____.______ W____.______ |
