## Supplementary material for "Development and Evaluation of a Clinical Guideline for a Pediatric Telemedicine and Medication Delivery Service: A Prospective Cohort Study in Haiti": S3 Medication Formulary

**Supplementary Text 3.**
Formulary: Version 5 28 2020

| Name | Formulation | Indication | Potential common side-effect | Dosage |
|---|---|---|---|---|
| **1 Non-opioids and non-steroidal anti-inflammatory medicines (NSAIM)** | | | | |
| Ibuprofen | **Oral liquid:** 200 mg/5 mL<br>**Tablet:** 200 mg; 400 mg; 600 mg | Inflamation / pain / fever | Stomach irritation | Children 10 mg/kg per dose by mouth every 6 hours as needed. Adults: 400 mg by mouth every 6 hours as needed. |
| Paracetamol | **Oral liquid:** 125 mg/5 mL<br>**Tablet:** 100 mg to 500 mg | Fever / pain / inflamation | No common side effect | Children: 15 mg/kg per dose by mouth every 6 hours as needed. Adults: 500 mg by mouth every 6 hours as needed. |
| **2. ANTIALLERGICS AND MEDICINES USED IN ANAPHYLAXIS** | | | | |
| Loratadine | **Oral liquid:** 1 mg/mL<br>**Tablet:** 10 mg | Mild allergy / allergic reaction | Mild tiredness | 2-<5 years old: 5 mg by mouth once daily.<br>5-18 years old: 10 mg by mouth once daily.<br>Adults: 20 mg by mouth once daily. |
| Diphenhydramine HCl | **Oral liquid:** 12.5mg/5mL<br>**Tablet:** 25 mg | Mild allergy / allergic reaction | Moderate tiredness | 2-5 years old: 6.25 mg by mouth every 6 hours as needed for allergic reaction.<br>5-10 years old: 12.5 mg by mouth every 6 hours as needed for allergic reaction.<br>Over 10 years: 25 mg by mouth every 6 hours as needed for allergic reaction. |
| **3. Anti-bacterials** | | | | |
| ***3.1 Beta-lactam medicines*** | | | | |
| Amoxicillin | **Powder for oral liquid:** 125 mg (as trihydrate)/5 mL; 250 mg (as trihydrate)/5 mL | Ear, lung, systemic infection | Rash/ allergic reaction | Children: 40 mg/kg per dose by mouth twice daily for 5 days. Adults: 875 mg by mouth every 12 hours for 5 days. |
| Amoxicillin / clavulanic acid | **Powder for oral liquid:** ___ mg Amox / ____ Clavulanic Acid per 5 mL | Lung infection when amoxicllin fails or is not available, or there is a UTI plus lung infection. | Rash/ allergic reaction | Children: 20 mg/kg every 8 hours for 10 days. Dose based on amoxicillin component.<br>Adults: 250 mg by mouth every 8 hours for 5 days. |
| Azithromycin | **Tablet:** 500 mg | Bloody diarrhea, watery diarrhea concerning for cholera, respiratory infection. | Rash/ allergic reaction | **For bloody diarrhea:** children 10 mg / kg by mouth once a day for 5 days (max 500 mg); adults 500 mg by mouth once a day for 5 days.<br>**For watery diarrhea**: chilldren 20 mg / kg by mouth as a single dose; adults (>15 yrs or >50kg) 1000 mg by mouth as a single dose.<br>**Bacterial pneumonia** (primary or secondary): children 10 mg/kg (max 500mg) per dose by mouth once, then 5 mg/kg (max 250mg) by mouth daily for days 2-5; adults 500mg by mouth once, then 250 mg by mouth daily for days 2-5. |
| Cefalexin (Cephalexin) | **Powder for reconstitution with water:** 125 mg/5 mL; 250 mg/5 mL (anhydrous).<br>**Solid oral dosage form:** 250 mg (as monohydrate). | Skin infection, urinary tract infection | Rash/ allergic reaction | **Skin Infection:** children 25 mg/kg per dose by mouth twice a day for five days; adults adults 500 mg every 12 hours for 5 days.<br>**Urinary tract infection:** children 25 mg/kg per dose by mouth four times a day for 5 days; adults 500 mg twice a day for 5 days. |
| ***3.2 Other antibacterials*** | | | | |
| Ciprofloxacin | **Tablet:** 500 mg | Bloody diarrhea, urinary tract infection | Rash / allergic reaction. Tendonitis. | **For bloody diarrhea:** children 15 mg/kg by mouth twice a day for three days; adults 500 mg by mouth twice a day for three days. |
| Doxycycline | **Tablet:** 100 mg | Infection, including watery diarrhea concerning for cholera | Rash / allergic reaction | **For watery diarrhea (cholera):** chilren 4 mg/kg mg by mouth once; adults 300 mg by mouth once. |
| Erythromycin | **Powder for oral liquid:** 125 mg/5 mL (as stearate or estolate or ethyl succinate)<br>**Solid oral dosage form:** 250 mg (as stearate or estolate or ethyl succinate) | Infection, including watery diarrhea concerning for cholera<br>Infection, including watery diarrhea concerning for cholera | Rash / allergic reaction<br>Rash / allergic reaction | **For watery diarrhea (cholera) for children less than 8 years**: Erythromycin 12.5 mg / kg per dose by mouth four times a day for 3 days; maximum single dose is 500 mg |
| Levofloxacin | **Tablet:** 500 mg | Moderate to severe primary / secondary bacterial pneumonia (COVID associated). | Rash / allergic reaction | **For pneumonia:** Children 10 mg/kg by mouth once daily by mouth for 7 days. Adults 750 mg by mouth once daily for 5 days. |
| Sulfamethoxazole + trimethoprim | **Oral liquid:** 200 mg + 40 mg/5 mL<br>**Tablet:** 100 mg + 20 mg; 400 mg + 80 mg | Infection, including skin and urinary tract infection (UTI). | Rash / allergic reaction. Avoid in children less than 2 months. | **UTI:** Children 10 mg/kg trimethoprim per dose by mouth twice a day for 5 days (WHO); adults sulfamethoxazole 800 mg/trimethoprim 160 mg orally twice daily for 3 days.<br>**Skin infection/ abscess**: children 4 mg/kg trimethoprim per dose by mouth twice a day for 5 days; adults sulfamethoxazole 800 mg/trimethoprim 160 mg orally twice daily for 5 days.(Doses based on the trimethoprim component) |
| **4. Anti-fungal medicines** | | | | |
| Nystatin (oral) | **Lozenge:** 100 000 IU<br>**Oral Liquid:** 50 mg/5mL; 100000 IU/mL | Oral thrush (fungus) | Rash / allergic reaction | Thrush: 2 ml by mouth four times a day until resolution |
| **5. DERMATOLOGICAL MEDICINES (topical)** | | | | |
| **6.1 Anti-infective medicines** | | | | |
| Antibiotic cream (neomycin sulfate, bacitracin zinc and polymyxin B) | **Cream** | Bacterial Skin infection | Rash / allergic reaction | Apply topically as needed 3-4 times daily |
| Miconazole (topical) | **Cream or ointment:** 2% (nitrate) | Fungal skin infection (diaper rash) | Rash / allergic reaction | Apply topically as needed 3-4 times daily |
| **6.2 Anti-inflammatory and anti-pruritic medicines** | | | | |
| Hydrocortisone (topical) | **Cream or ointment:** 1% (acetate) | Mild inflammation in skin. | None | Apply topically as needed 3-4 times daily |
| **6.3 Scabicides and pediculicides** | | | | |

| | | | | |
|---|---|---|---|---|
| Benzyl benzoate (topical) | **Liquid:** 15% | Insect irritation (lice, scabies) | Rash / allergic reaction | **Scabies:** Apply from hairline to toes once at bedtime. Rinse in morning. Repeat in 1 week. Infants: Dilute 1 part BB to 1 part water.<br>**Pediculosis (lice):** Apply cream to hair at bedtime. Rinse in the morning. Repeat in 1 week. |
| Ivermectin | **Oral liquid:** ___ mg /5 mL<br>**Tablet:** 6 mg | Scabies (severe) | Rash / allergic reaction | Children: 0.2 mg/kg per dose by mouth once. Repeat in two weeks. Only for children that weigh at least 15 kg. Take with food. |
| Permethrin (topical) | **Cream:** 5%<br>**Lotion**: 1% | Insect irritation (lice, scabies) | Rash / allergic reaction | **Scabies**: Apply 5% cream from hairline to toes once at bedtime. Rinse in morning. Repeat in 1 week.<br>**Pediculosis (lice):** Apply 1% cream to hair. Rinse after 1 hour. Repeat in 1 week.<br><br>Wash bedding and clothes with hot water. |
| **7. Fluids and medicines used in diarrhoea** | | | | |
| ***7.1 Oral rehydration*** | | | | |
| Oral rehydration solution | **Powder for dilution** in 200 mL; 500 mL; 1 L | Rehydration / Hydration | None | See guidelines |
| ***7.2 Medicines for diarrhoea*** | | | | |
| Zinc sulfate | **Solid oral dosage:** 20 mg | Shortens diarrhea course | None | 2 mo - <6 mo: 10 mg by mouth once daily for 10 days.<br>6 mo - <5 years: 20 mg by mouth once daily for 10 days. |
| **8.1 Anti-asthmatic medicines** | | | | |
| Salbutamol | **Metered dose inhaler (aerosol):** 100 micrograms (as sulfate) per dose<br>**Respirator solution for use in nebulizers:** 5 mg (as sulfate)/mL | Asthma/ reactive airway disease. | Increases heart rate. Tremor. | 3 years and older: Two puffs (100 ug/puff) with a spacer every 4 hours for 5 days for wheeze. Under three years with yellow sign and wheeze need hospital referral; green sign and wheeze need morning clinic follow-up. |
| **8.1 Anti-emetic** | | | | |
| Ondansetron | **Oral dissolvable tab (ODT):** 4mg or 8 mg<br>**Oral liquid:** 4 mg/5 ml | Nause | None | 1-<4 years: 2 mg by mouth once<br>≥4 years: 4 mg by mouth once |
