## Supplementary material for "Development and Evaluation of a Clinical Guideline for a Pediatric Telemedicine and Medication Delivery Service: A Prospective Cohort Study in Haiti": STable 1

Table S1. Performance of components of the dehydration screen and assessment during the virtual exam

| Component | Description | Total | CC+ HH+ (%)[a] | CC+ HH- (%)[a] | CC- HH+ (%)[a] | CC- HH- (%)[a] | Sens. (95% CI) | Spec. (95% CI) | PPV (95% CI) | NPV (95% CI) | kappa (95% CI) |
|---|---|---|---|---|---|---|---|---|---|---|---|
| Dehydration screen | | | | | | | | | | | |
| Urinated | | 318 | 316 (99) | 1 (<1) | 0 | 1 (<1) | 100 | 50 (0-100) | 100 (99-100) | 100 | 0.67 (0.05-1.00) |
| Tears | | 249 | 246 (99) | 0 | 2 (1) | 1 (<1) | 99 (98-100) | 100 | 100 | 33 (0-87) | 0.50 (-0.10-1.00) |
| Dehydration assessment | | | | | | | | | | | |
| Disposition ≥5 yrs | Alert ≥5 yrs | 11 | 11 (100) | 0 | 0 | 0 | 100 | --- | 100 | --- | --- |
| Disposition <5 yrs | | | | | | | | | | | |
| | Alert <5 yrs | 86 | 80 (93) | 2 (2) | 3 (4) | 1 (1) | 96 (92-100) | 33.3 (0-86.7) | 98 (95-99) | 25 (5-70) | 0.26 (-0.19-0.70) |
| | Irritable | 86 | 1 (1) | 3 (4) | 2 (2) | 80 (93) | 33 (0-87) | 96.4 (92.4-100) | 25 (5-70) | 98 (95-99) | 0.26 (-0.19-0.70) |
| | Lethargic | 86 | 0 | 0 | 0 | 86 (100) | --- | 100 | --- | 100 | --- |
| Eyes | Sunken eyes | 96 | 2 (2) | 2 (2) | 2 (2) | 90 (94) | 50 (1-99) | 97.8 (94.9-100) | 50 (16-84) | 98 (94-99) | 0.48-(0.04-0.92) |
| Thirst | Normal | 97 | 87 (90) | 1 (1) | 3 (3) | 6 (6) | 97 (93-100) | 86 (60-100) | 99 (93-100) | 67 (39-86) | 0.73 (0.48-0.98) |
| | Thirsty | 97 | 6 (6) | 3 (3) | 1 (1) | 87 (90) | 86 (60-100) | 97 (93-100) | 67 (39-86) | 67 (39-86) | 0.73 (0.48-0.98) |
| | Unable to drink | 97 | 0 | 0 | 0 | 97 (100) | --- | 100 | --- | 100 | --- |
| Skin pinch | Normal | 96 | 93 (97) | 1 (1) | 2 (2) | 0 | 98 (95-100) | 0 | 99 (99-99) | 0 | -0.01 (-0.03-0.01) |
| | Slow | 96 | 0 | 2 (2) | 1 (1) | 93 (97) | 0 | 97.9 (95.0-100) | 0 | 99 (99-99) | -0.01(-0.03-0.01) |
| | Very slow | 96 | 0 | 0 | 0 | 96 (100) | --- | 100 | --- | 100 | --- |
| Dehydration severity | | | | | | | | | | | |
| | None | 97 | 88 (91) | 1 (1) | 3 (3) | 5 (5) | 97 (93-100) | 83 (53-100) | 99 (97-100) | 63 (29-96) | 0.69 (0.41-0.98) |
| | Moderate | 97 | 5 (5) | 3 (3) | 1 (1) | 88 (91) | 83 (54-100) | 97 (93-100) | 63-(29-96) | 99 (97-100) | 0.69 (0.41-0.98) |

[a] CC= call center, HH= household, '+' = present, '-' = absent.
